# Effect of reduction in brain amyloid levels on change in cognitive and functional decline in randomized clinical trials: an updated instrumental variable meta-analysis

**DOI:** 10.1101/2022.04.01.22273253

**Authors:** Menglan Pang, Ling Zhu, Audrey Gabelle, Arie R. Gafson, Robert W Platt, James E Galvin, Pierre Krolak-Salmon, Ivana Rubino, Carl de Moor, Shibeshih Belachew, Changyu Shen

**Author notes:** Corresponding author: Changyu Shen, PhD, Address: 225 Binney St, Cambridge, MA 02142, USA.

## Abstract

**Objective:** To update a recently published analysis exploring the causal association between positron emission tomography (PET)-measured change in brain β-amyloid plaque and cognitive decline in patients with Alzheimer’s disease (AD) enrolled in randomized clinical trials (RCTs).

**Design:** Updated instrumental variable meta-analysis.

**Setting:** Sixteen RCTs were included in this updated meta-analysis versus 14 in the original publication by Ackley *et al*.^1^ Data sources were ClinicalTrials.org, Alzheimer Research Forum (alzforum.org), PubMed and clinical study reports from 2015 to March 1, 2022. Three researchers extracted data from the data sources independently and subsequently resolved any discrepancy.

**Population:** RCTs that evaluated β-amyloid targeting therapies and enrolled adult patients with AD dementia or mild cognitive impairment due to AD with data on β-amyloid as measured by PET and clinical outcome measures.

**Main outcome measures:** An instrumental variable meta-analysis was performed to compute trial and drug-specific estimates and pooled estimates of the effect of change in PET β-amyloid standard uptake value ratio (SUVR) on cognitive and functional decline with 95% confidence intervals (CIs) and associated p-values. This analysis updated and expanded a prior meta-analysis by Ackley *et al*.^1^ using the same methodology and clinical outcome measures: Clinical Dementia Rating–Sum of Boxes (CDR-SB), Alzheimer’s Disease Assessment Scale–Cognitive Subscale (ADAS-Cog), and Mini-Mental State Examination (MMSE).

**Results:** The reduction of PET-measured β-amyloid induced a statistically significant reduction in cognitive and functional decline. The effect size was characterized by an estimated change (95% CI) of 0.09 (0.034, 0.15) on the CDR-SB; 0.33 (0.12, 0.55) on the ADAS-Cog; and 0.13 (0.017, 0.24) on the MMSE for each decrease of 0.1-unit in PET β-amyloid SUVR.

**Conclusion:** This updated instrumental variable meta-analysis of 16 RCTs provides statistically significant evidence of a causal relationship between the reduction in brain β-amyloid plaque and the reduction in cognitive and functional decline in patients with Alzheimer’s disease.

## INTRODUCTION

Alzheimer’s disease (AD) is defined pathologically by the presence of β-amyloid deposits, tau-containing neurofibrillary tangles, neuronal injury and degeneration.^2^ Clinically, the disease course is characterized by progressive cognitive decline, behavioral changes, increased dependency in activities of daily living and increased burden for caregivers and society.^3 4^ Genetic data, preclinical models, biomarkers, and clinical observations^5–8^ have highlighted that removal of brain parenchymal β-amyloid remains a relevant target for slowing disease progression. Nonetheless, randomized clinical trials (RCTs) of drugs that target the reduction in production or removal of β-amyloid have generated inconsistent results, and the link between change in β-amyloid pathology and cognitive/functional performance is hotly debated.^9^

One possible reason for the inconsistent evidence that β-amyloid is a clinically valid target in AD may be that patient-level correlation analyses between change in β-amyloid and change in cognition using data from a single trial may have insufficient statistical power and/or confounding bias. To address these limitations, Ackley *et al*.^1^ published a meta-analysis using an instrumental variable approach that integrated 14 RCTs with data available up until April 30, 2020. The RCTs were selected using the Alzheimer Research Forum (alzforum.org) and from ClinicalTrials.gov and required quantitative measurements of change in a cognitive score and brain β-amyloid data using the standardized uptake value ratio (SUVR) obtained from β-amyloid positron emission tomography (PET). The authors then used randomization to treatment groups as the instrumental variable to eliminate confounding bias, permitting valid causal inference on the effect of average PET-measured β-amyloid reduction on average cognitive and functional decline within and across multiple studies. In the Ackley *et al*. study,^1^ the pooled estimates changes were 0.058 (95% CI: −0.031 to 0.15) and 0.034 (95% CI: −0.056 to 0.12) on the Clinical Dementia Rating Scale–Sum of Boxes (CDR-SB) and Mini-Mental State Examination (MMSE) scores, respectively, for each 0.1-unit reduction in the PET β-amyloid SUVR.

In their original work, Ackley *et al*.^1^ provided a publicly available web interface containing an interactive version of their analytic approach to encourage recalculation of their results when updated or new data became available. The authors also acknowledged that they did not have full access to the source data for certain trials, and potential errors in input data might have occurred in their analysis. In response to the invitation to the research community, we reviewed the data included in their meta-analysis and identified several inconsistencies and some limitations associated with the data retrieval process for the 14 RCTs. In addition, there were trials not included in the initial analysis, partially due to data not being available in the public domain at the time of publication, which could potentially improve the accuracy of the pooled estimate. In this article, we provided an updated version of this instrumental variable meta-analysis by performing several data updates and by adding data from two additional RCTs. Our updated analysis demonstrates that a reduction in β-amyloid plaque is causally and consistently associated with a statistically significant reduction in cognitive and functional decline as measured by changes in the CDR-SB, Alzheimer’s Disease Assessment Scale–Cognitive Subscale (ADAS-Cog), and MMSE.

## METHODS

For trials included in the initial publication by Ackley *et al*.^1^, three authors independently reviewed the source data for β-amyloid PET SUVR, CDR-SB, ADAS-Cog and MMSE from clinicaltrials.gov, published articles and clinical study reports, and subsequently resolved any discrepancy. In addition, one author applied the same search strategy and selection criteria as the original paper^1^ to seek supplemental trial data eligible to be included in this updated analysis, except that trials were further required to have a “Last Updated Posted” date at clinicaltrials.gov between 30 April 2020 (the date of data cut of the initial publication) and 1 March 2022.

### Updated data from the original analysis

For 12 of the 14 studies for which Ackley *et al*. retrieved the data entirely from the original source (ClinicalTrials.gov, peer reviewed publications, or other publicly available materials), we identified and eliminated several inconsistencies between the data used by Ackley *et al*. and the source data (**Table A1 in the Appendix 1**). For example, the inputted standard errors (SEs) for SUVR and MMSE measures from four of these trials were inconsistent with the data source, including standard deviations (SDs) of the SUVR being used in place of the SEs in two trials (bapineuzumab-1 [<u>NCT00575055</u>] and gantenerumab (Scarlet Road) [<u>NCT01224106</u>]). In addition, data for the change in CDR-SB and ADAS-Cog in solanezumab 1&2^10^ (solanezumab-1&2 included data from an integrated population of mild AD from both solanezumab (EXPEDITION) [<u>NCT00905372</u>] and solanezumab (EXPEDITION2) [<u>NCT00904683</u>]) were incorrectly reversed between the treatment and placebo groups. Finally, the original source data of aducanumab-1 (EMERGE) [<u>NCT02484547</u>] and aducanumab-2 (ENGAGE) [<u>NCT02477800</u>] trials were based on partial data, which were updated with the latest data from ClinicalTrials.gov.

For the other two RCTs, Ackley e*t al*. obtained at least part of the data through estimating algorithms rather than directly from the source data (**Table A2 in the Appendix 1**). Specifically, MMSE changes in the lecanemab [<u>NCT01767311</u>] and verubecestat-2 (APECS) [<u>NCT01953601</u>] trials were estimated based on the changes in the Alzheimer’s Disease Composite Score (ADCOMS) and CDR-SB, respectively. We updated MMSE change scores using source data from a clinical study report for lecanemab and a publication for verubecestat-2 (APECS).^11^ We also identified and corrected an inconsistency in the SE values for verubecestat-2 (APECS) PET β-amyloid SUVR in the original publication (**Table A1 in the Appendix 1**).

### Inclusion of two additional RCTs

We expanded the initial meta-analysis by including two additional RCTs of β-amyloid targeting drugs (aducanumab-3 (PRIME) [<u>NCT01677572</u>] and donanemab (TRAILBLAZER-ALZ) [<u>NCT03367403</u>]), with data on change in PET β-amyloid SUVR and clinical outcome measures at week 54 for PRIME and week 76 for donanemab (TRAILBLAZER-ALZ) available in the public domain.^12^

### Statistical analysis

A program in the *R* computation environment (**Appendix 2**) was written based on the same methodology described in the original publication^1^ and replicated the output from the publicly available web interface. A new program was written because the publicly available web interface only permitted the inclusion of one additional RCT. We computed both trial- and drug-specific and pooled estimates of the effect of a reduction in PET β-amyloid SUVR on the change of CDR-SB, ADAS-Cog, and MMSE together with 95% confidence intervals (CIs) and the associated p-values. We used the same representation of the effect estimates as in the original meta-analysis, where it was defined as the change in clinical endpoints per 0.1-unit change in PET β-amyloid SUVR (a 0.1-unit reduction in PET β-amyloid SUVR is a mathematical representation to measure the effect of a continuous exposure where the choice of unit change is driven by convenience; it does not confer to this arbitrary unit any actual clinical meaningfulness).

Solanezumab-1&2 (EXPEDITION EXT) (<u>NCT01127633</u>) was an extension of solanezumab-1&2 with a longer follow-up period for continued safety monitoring. In order to maximize the follow-up period and similarly to the original study,^1^ we included the data from solanezumab-1&2 (EXPEDITION EXT) (rather than from solanezumab-1&2 *and* solanezumab-1&2 (EXPEDITION EXT)) in all pooled estimates as well as the drug-specific estimate for solanezumab. For ADAS-Cog, the aducanumab-3 (PRIME) trial was not included in the pooled estimates nor in the drug-specific estimate because this endpoint was not measured in this trial. Finally, Bexarotene (BEAT-AD) [<u>NCT01782742</u>] was not included in the pooled estimate for CDR-SB as no CDR-SB source data were available.

We performed similar sensitivity analyses as in Ackley *et al*.^1^ by (1) including all published data, (2) only including all antibody data, and (3) only including all *published* antibody data. Note that lecanemab was the only trial with unpublished data in our analyses, whereas in the original manuscript, both lecanemab and the two aducanumab trials (EMERGE and ENGAGE) relied on unpublished data. We also performed the following additional sensitivity analyses:

1. Included only the most recent β-amyloid targeting antibodies, i.e. gantenerumab (Scarlet Road), aducanumab (EMERGE, ENGAGE, and PRIME), lecanemab, and donanemab (TRAILBLAZER-ALZ).
2. Excluded the two RCTs evaluating verubecestat (EPOCH & APECS) [<u>NCT01739348 & NCT01953601</u>] as evidence of treatment-associated cognitive worsening^13^ due to the inhibition of β-secretase 1 which may indicate off-target effects.
3. Applied data updates (termed “*initial trials with data updates*” in **Figure 1**) as described in the methods without including the two new additional RCTs data sets (aducanumab-3 (PRIME) and donanemab (TRAILBLAZER-ALZ)).
4. Excluded the lecanemab trial month 12 data to address the limitation of not accounting for the correlation between data from week 53 (month 12) and week 79 (month 18).
5. Excluded donanemab (TRAILBLAZER-ALZ) trial data, because the SUVR values in this trial were converted from the Centiloid scale using an equation.^14^ This sensitivity analysis was performed to eliminate potential inaccuracies resulting from this conversion.
6. Excluded both lecanemab trial month 12 and donanemab (TRAILBLAZER-ALZ) trial data for reasons specified in (d) and (e).

**Figure 1.**
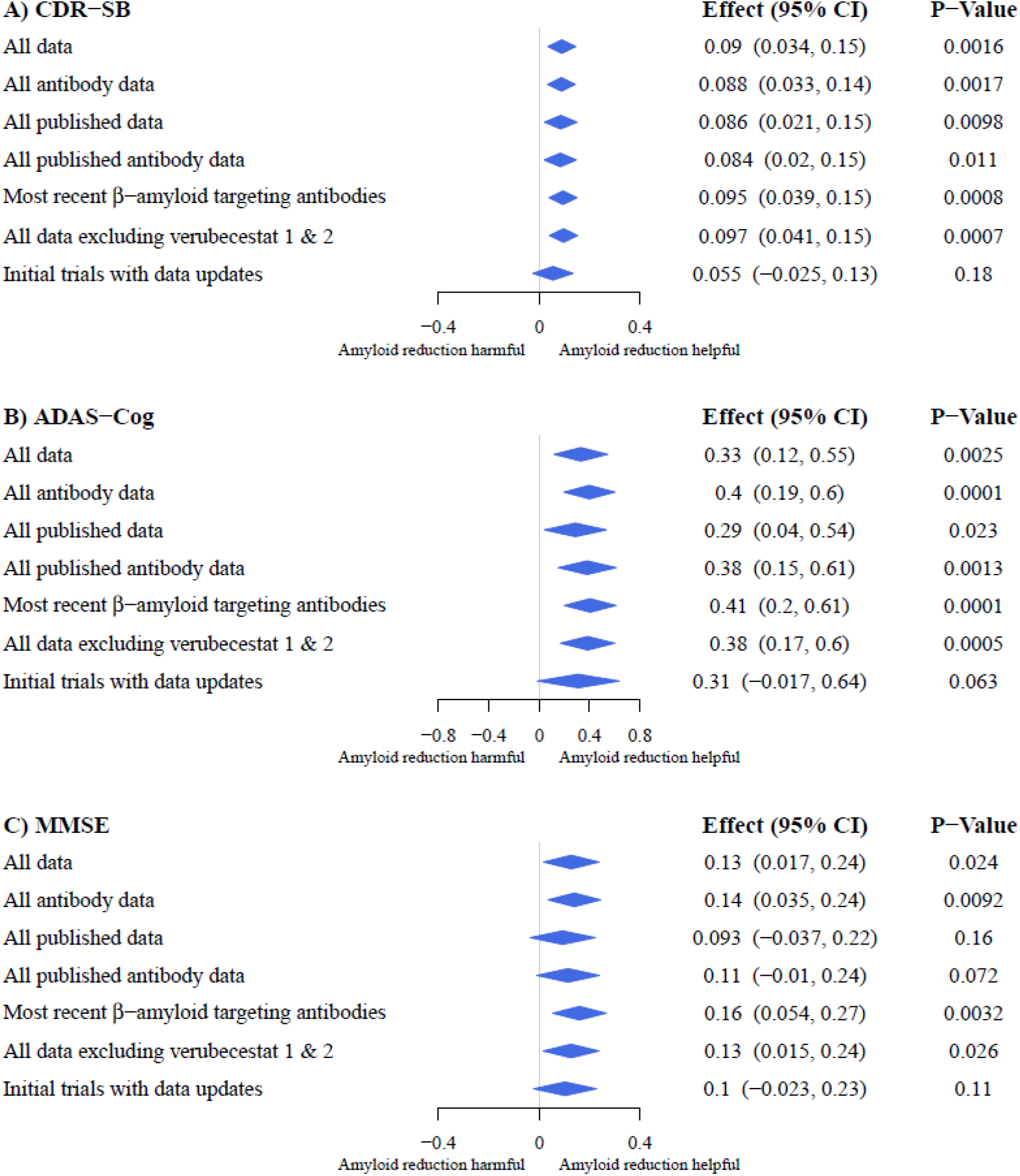
Forest plots of the pooled estimates representing the effect of a decrease in brain β-amyloid as measured by PET on change in A) CDR-SB, B) ADAS-Cog, and C) MMSE. Positive values of the effect estimate—for each 0.1-unit reduction in PET SUVR—indicate that β-amyloid reduction reduces cognitive decline. Center and width of diamonds represent pooled estimates and 95% confidence intervals, respectively. The trial of lecanemab is unpublished and was excluded from the “*All published data*” and “*All published antibody data*” categories in the sensitivity analyses. Most recent β-amyloid targeting antibodies included gantenerumab, aducanumab (ENGAGE, EMERGE, PRIME), lecanemab, and donanemab trials. ADAS-Cog, Alzheimer’s Disease Assessment Scale– Cognitive Subscale; CDR-SB, Clinical Dementia Rating–Sum of Boxes; CI, confidence interval; MMSE, Mini-Mental State Examination.

## RESULTS

For the pooled estimates on “*All Data*,” this updated meta-analysis showed statistically significant evidence of a causal relationship between the reduction in brain β-amyloid plaque levels and reduction in cognitive and functional decline as measured by CDR-SB (0.09 point per each 0.1-unit reduction in PET β-amyloid SUVR; 95% CI: 0.034, 0.15; with a p-value of 0.0016) (**Figure 1A**). The updated point estimate is approximately 1.5 times the original estimate (0.058 with 95% CI: −0.031, 0.15) (**Table 1**).^1^ Multiple sensitivity analyses were performed to exclude the influence of specific trials affected by inherent limitations as described in the Methods. These sensitivity analyses (**Figure 1A and Figure A2 in the Appendix 3**) yielded similar point estimates for CDR-SB, with p-values < 0.05, except for “*initial trials with data updates*.”

**Table 1.** Comparison of the main results between the original and updated analysis on the effect of a reduction in β-amyloid SUVR on change in cognitive endpoints. Positive values of the effect estimate—for each 0.1-unit reduction in PET SUVR—indicate that β-amyloid reduction slows cognitive decline.

| Cognitive endpoints | Original analysis<br>("All data") | Updated analysis<br>("All data") |  |
| --- | --- | --- | --- |
|  | Effect (95% CI) | Effect (95% CI) | P-Value |
| CDR-SB | 0.058 (−0.031, 0.15) | 0.09 (0.034, 0.15) | 0.0016 |
| ADAS-Cog | NA | 0.33 (0.12, 0.55) | 0.0025 |
| MMSE | 0.034 (−0.056, 0.12) | 0.13 (0.017, 0.24) | 0.024 |

ADAS-Cog, Alzheimer’s Disease Assessment Scale–Cognitive Subscale; CDR-SB, Clinical Dementia Rating–Sum of Boxes; CI, confidence interval; MMSE, Mini-Mental State Examination; NA, not available.

The updated meta-analysis further showed a statistically significant causal effect of β-amyloid reduction on reduction in cognitive decline as measured by ADAS-Cog (0.33 point per each 0.1-unit reduction in PET β-amyloid SUVR; 95% CI: 0.12, 0.55; with a p-value of 0.0025) (**Figure 1B**), which remained the case in all sensitivity analyses performed (**Figure 1B** and **Figure A2 in the Appendix 3**) except for *“initial trials with data updates*.*”* We were not able to compare the updated pooled estimate with the original estimate for ADAS-Cog as this was not provided in the original work from Ackley *et al*.^1^

The updated meta-analysis also showed a statistically significant causal relationship between β-amyloid reduction and reduced decline as measured by the MMSE (0.13 point per each 0.1-unit reduction in PET β-amyloid SUVR; 95% CI: 0.017, 0.24; with a p-value of 0.024) (**Figure 1C**). The pooled estimate in the updated analysis was four times the original estimate (0.034 with 95% CI: −0.056, 0.12) (**Table 1**).^1^ When we performed only data updates as described in the Methods without including the two new additional RCTs data sets (aducanumab-3 [PRIME] and donanemab [TRAILBLAZER-ALZ]), the updated meta-analysis also demonstrated a substantial shift from the original results on the MMSE endpoint (0.10 point per each 0.1-unit reduction in PET β-amyloid SUVR; 95% CI: −0.023, 0.23), although this was not statistically significant.

Finally, a sensitivity analysis including only the most recent β-amyloid targeting antibodies (i.e. gantenerumab, aducanumab, lecanemab, and donanemab), showed overall numerically stronger effects across all three endpoints: CDR-SB (0.095 point per each 0.1-unit reduction in PET β-amyloid SUVR; 95% CI: 0.039, 0.15; with a p-value of 0.0008), ADAS-Cog (0.41 point per each 0.1-unit reduction in PET β-amyloid SUVR; 95% CI: 0.2, 0.61; with a p-value of 0.0001), and MMSE (0.16 point per each 0.1-unit reduction in PET β-amyloid SUVR; 95% CI: 0.054, 0.27; with a p-value of 0.0032) with consistent statistical significance (**Figure 1**).

Trial- and drug-specific estimates for CDR-SB, ADAS-Cog, and MMSE are presented in **Figure A1 in the Appendix 3**. The RCTs included in this updated analysis are summarized in **Appendix 4**.

## DISCUSSION

In this study, we updated a previously published instrumental variable meta-analysis^1^ of the effect of a reduction in β-amyloid plaque as measured by PET on change in cognitive decline, by correcting and improving the original data inputs and increasing the number of RCTs included. In contrast to the initial analysis, our study demonstrates that a reduction in β-amyloid is causally and consistently associated with a statistically significant reduction in cognitive and functional decline as measured by changes in CDR-SB, ADAS-Cog, and MMSE.

### Strengths and limitations

A unique strength of the methodology developed in the original publication^1^ is the use of the instrumental variable approach to address the issue of potential confounding bias found in correlation analysis. In addition, the integration of data from multiple trials substantially improves statistical precision relative to any individual trial.

Despite being categorized as level 1 evidence with multiple advantages, meta-analyses are not without limitations,^15^ some of which have already been highlighted in the original publication.^1 16^ Additional points of caution in interpretation should be emphasized.

First, subjects included in this meta-analysis are at different stages of AD, from mild cognitive impairment due to AD to moderate stage of AD dementia. In addition, β-amyloid positivity (measured with visual scale or PET SUVR threshold) was not a prerequisite inclusion criterion in the majority of included studies (9 of 16 studies). It is biologically plausible that subjects in this heterogeneous population may derive different levels of benefit in cognitive function from β-amyloid reduction. In this context, combining those individuals with prodromal, mild, or moderate AD, in which the underlying pathology (β-amyloid load and downstream processes) may vary, could cause difficulty in interpreting the results.^16^

Second, while we assume a linearity between β-amyloid deposition and cognitive and functional decline, the precise temporal and spatial dynamic of this relationship is not well understood. For example, although a change in SUVR may be a generalizable metric, it may not optimally reflect spatially and temporally heterogenous properties of the underlying pathology associated with and relevant to AD and its clinical manifestations, particularly at the early stage of the disease. As such, recent findings using regional SUVR data have demonstrated promise.^17 18^

Third, the instrumental variable analysis requires the absence of off-target effects from the therapeutic agents. In our present study, we partially addressed this limitation by performing a sensitivity analysis including only trials with antibody therapies that, due to their specificity and direct impact on β-amyloid, may provide more accurate estimates of this relationship. However, this would not have addressed the highly complex temporal relationship that likely exists between the impact of amyloid plaque removal and the decline in cognition and function that is inherently limited by the short-term setting of most typical clinical trials and where a potentially delayed-onset effect cannot be captured.

Fourth, the various radiotracers used in trials included in this analysis differ in their sensitivity, specificity, variability, dynamic ranges, and other performance metrics ^19–23^. Therefore, although different amyloid tracers produce highly consistent and highly correlated results, SUVR derived from different tracers are not equivalent and should be compared or combined with some caution. Furthermore, although amyloid PET has been shown to be robust against different quantification methods ^24^, different analysis pipelines may also have resulted in small differences between studies.

Finally, the assessment of change in AD-related impairment of cognition and function at the early phases of AD is not fully addressed by the clinical scores utilized in these clinical trials, in particular the MMSE, due to its limited sensitivity to capture progression in cognitive decline. This was further emphasized in our current study by the demonstration of higher p-values for MMSE compared with CDR-SB and ADAS-Cog endpoints **(Figure 1)**.

### Implications

That a statistically significant causal association between decreasing PET-measured β-amyloid plaque and reduction of cognitive and functional decline was consistently demonstrated on all three clinical endpoints despite some of the aforementioned limitations highlights the potential of β-amyloid as a viable biological target in AD. As more data accumulate in the future, we hope that some of the limitations can be addressed by analysis of data with longer follow-up periods. In addition, the ability to harmonize the data on β-amyloid load by using Centiloids will further account for the different properties of β-amyloid radiotracers. These efforts will shed more light on the effect of β-amyloid reduction on cognitive and functional decline and guide future drug development and clinical applications.

## CONCLUSIONS

This updated instrumental variable meta-analysis, which is based on 16 randomized clinical trials and expands on the original work by Ackley *et al*.,^1^ demonstrated a consistent and statistically significant impact of PET β-amyloid SUVR change on three commonly used clinical outcome measures. More data and research are needed to further characterize the precise spatial/temporal properties and potential heterogeneity of the causative relationship between β-amyloid clearance and cognitive and functional trajectory within the AD continuum.

## Data Availability

All data produced in the present work are contained in the manuscript

## PATIENT AND PUBLIC INVOLVEMENT

Patients or the public were not involved in the design, or conduct, or reporting, or dissemination plans of this research.

## ACKNOWLEDGMENTS

We would like to thank Tammy Jiang for quality-checking the input data; John O’Gorman, Samantha Budd Haeberlein, Tucker Ward, Kathleen Grobben, Annie Racine and Priya Singhal for reviewing the manuscript and providing helpful comments.

