## Appendix for "Effect of reduction in brain amyloid levels on change in cognitive and functional decline in randomized clinical trials: an updated instrumental variable meta-analysis"

### Appendices

#### Appendix 1: Details of data quality improvement and results comparison

**Table A1. Summary of data inconsistency between inputted data presented in Ackley *et al.*[1] and source data; data inconsistency was corrected to generate the updated results.**

| Trial | Measure | Statistics | Description of data inconsistency | Effect (95% CI) | | |
|---|---|---|---|---|---|---|
| | | | | Clinical endpoint | Original results | Updated results |
| Bapineuzumab-1 | SUVR | SE of change | SD was used as SE | MMSE | −1 (−2.8, 0.75) | −1 (−2.4, 0.35) |
| Solanezumab-1&2 (EXPEDITION & EXPEDITION2) | SUVR | SE of change | Values were switched between the treatment and placebo group | | 3.1 (−2.4, 8.6) | 3.1 (−2.4, 8.6) |
| | MMSE | N | | | | |
| Gantenerumab (Scarlet Road) | SUVR | SE of change | SD was used as SE | | **0.26 (−1, 1.5)** | **0.31 (−0.61, 1.2)** |
| Verubecestat-2 (APECS) | SUVR | SE of change | Values were incorrect for unknown reasons | | −0.59 (−1.7, 0.52) | −0.6 (−1.6, 0.44) |
| Aducanumab-2 (ENGAGE) | MMSE | SE of change | Values were incorrectly extracted from graph | | −0.014 **(−0.13, 0.1)** | −0.011 **(−0.25, 0.23)** |
| Solanezumab-1&2 (EXPEDITION & EXPEDITION2) | CDR-SB | Mean change | Values were switched between the treatment and placebo group | CDR-SB | **−0.53** (−2, 0.95) | **0.53** (−0.95, 2) |
| | ADAS-Cog | Mean change | | ADAS-Cog | **−7.1** (−20, 5.6) | **7.1** (−5.5, 20) |

ADAS-Cog, Alzheimer's Disease Assessment Scale–Cognitive Subscale; CDR-SB, Clinical Dementia Rating–Sum of Boxes; CI, confidence interval; MMSE, Mini-Mental State Examination; PET, positron emission tomography; SD, standard deviation; SE, standard error; SUVR, standard uptake value ratio. Effect represents change in clinical outcome measure for each 0.1 reduction in SUVR with positive value indicating beneficial effect of beta-amyloid reduction.

**Table A2. Summary of data conversion implemented in Ackley *et al.*[1]; direct source data were used to generate the updated results.**

| Trial | Measure | Statistics | Description of data conversion | Effect (95% CI) | | |
|---|---|---|---|---|---|---|
| | | | | Clinical endpoint | Original results | Updated results |
| Verubecestat-2 (APECS) | MMSE | Mean change & SE of change | Data were converted from CDR-SB | MMSE | −0.59 (−1.7, 0.52) | −0.6 (−1.6, 0.44) |
| Lecanemab | MMSE | Mean change & SE of change | Data were converted from ADCOMS | | 0.19 (−0.038, 0.42) | 0.2 (0.0057, 0.39) |

ADCOMS, Alzheimer's Disease Composite Score; CI, confidence interval; MMSE, Mini-Mental State Examination; SD, standard deviation; SE, standard error. Effect represents change in clinical outcome measure for each 0.1 reduction in SUVR with positive value indicating beneficial effect of beta-amyloid reduction.

### Major change in trial-specific results by correcting data inconsistency and using source data

For MMSE, we observed a major difference in the 95% CI between the updated result and the original result for aducanumab-2 (ENGAGE) (NCT02477800): (−0.25, 0.23) versus (−0.13, 0.1). The much narrower 95% CI in the original analysis is mainly due to an error in the SE of MMSE change (**0.1** was used in the original analysis instead of the actual value of **0.21**). For gantenerumab (Scarlet Road) [NCT01224106], the updated result yielded a larger point estimate with narrower 95% CI: 0.31 (−0.61, 1.2) versus 0.26 (−1, 1.5), because the SDs of change in the standard uptake value ratio were misused as SEs in the original inputted data, which overestimated the SEs by four times. For solanezumab-1&2 (EXPEDITION & EXPEDITION2), the original paper reported an estimate of −0.53 with 95% CI (−2, 0.95) for CDR-SB and −7.1 with 95% CI (−20, 5.6) for the Alzheimer's Disease Assessment Scale–Cognitive Subscale, which are the opposite of our updated analysis, because the original analysis reversed the data between the solanezumab and placebo arms.

For lecanemab [NCT01767311], our result based on direct MMSE source data provided a statistically significant estimate of 0.2 with 95% CI (0.0057, 0.39), whereas the original result based on data converted from ADCOMS reported an estimate of 0.19 (−0.038, 0.42), which was not statistically significant.

### Appendix 2: Details of statistical and computational methods

The same methodology via instrumental variable meta-analysis as proposed by the original paper was used to estimate the trial- and drug-specific effect as well as the pooled effect of amyloid reduction on cognitive decline. All statistically analysis was performed in *R* version 4.0.5. The maximum likelihood estimates were obtained using the "optim" function based on both "Nelder-Mead" and "BFGS" algorithms, and the estimate that achieves a larger likelihood was kept. For semagacestat-2 [NCT00594568] (referred to as LY450139-2 in Ackley *et al*. (PMID: 33632704)), the estimated effects on the Clinical Dementia Rating–Sum of Boxes and the Alzheimer's Disease Assessment Scale–Cognitive Subscale were unrealistically large based on the "Nelder-Mead" algorithm, therefore the results were based on the "BFGS" optimization although with a slightly smaller resulting likelihood.

For the donanemab (TRAILBLAZER-ALZ) (NCT03367403) trial, the difference in the standard uptake value ratio (SUVR) was converted from difference in Centiloid based on the following equation: SUVR = (Florbetapir Centiloids + 177)/183 [PMID: 30006100].

Without individual-level data, one cannot estimate and account for the covariance between the measured SUVR change and cognition change from the same cohort of subjects in the instrumental variable analysis. As such, the original analysis ignored the covariance. Nonetheless, this omission could cause inaccuracy in the estimation process. We performed analysis using individual-level data from aducanumab-1 (EMERGE) and aducanumab-2 (ENGAGE) to account for the covariance. The results were very similar to those obtained without accounting for the covariance. Therefore, the covariance was not accounted for in our updated analysis.

### Appendix 3: Trial- and drug-specific results and pooled results from sensitivity analyses

**Figure A1** A) Trial- and drug-specific estimates representing the effect of each 0.1-unit decrease in brain β-amyloid as measured by PET SUVR on change in CDR-SB in the clinical trials described alone and for all data from the verubecestat, solanezumab, bapineuzumab, and aducanumab drugs. CDR-SB, Clinical Dementia Rating-Sum of Boxes; CI, confidence interval; E/E, EMERGE and ENGAGE; E/E/P, EMERGE, ENGAGE and PRIME; PET, positron emission tomography; SUVR, standard uptake value ratio.

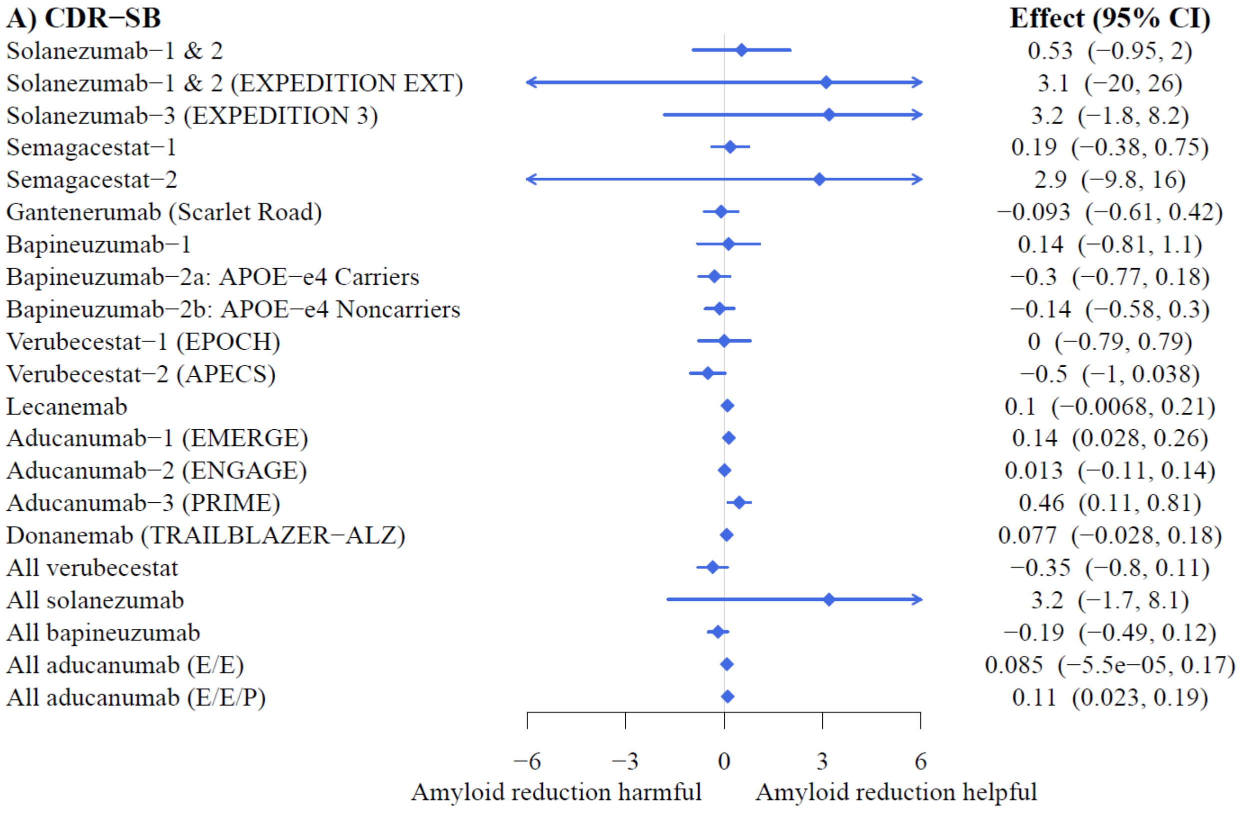

**Figure A1** – B) Trial- and drug-specific estimates representing the effect of each 0.1-unit decrease in brain β-amyloid as measured by PET SUVR on change in ADAS-Cog in the clinical trials described alone and for all data from the verubecestat, solanezumab, bapineuzumab, and aducanumab drugs. The version of the ADAS-Cog is provided in parentheses. ADAS-Cog, Alzheimer's Disease Assessment Scale–Cognitive Subscale; CI, confidence interval; E/E, EMERGE and ENGAGE; PET, positron emission tomography; SUVR, standard uptake value ratio.

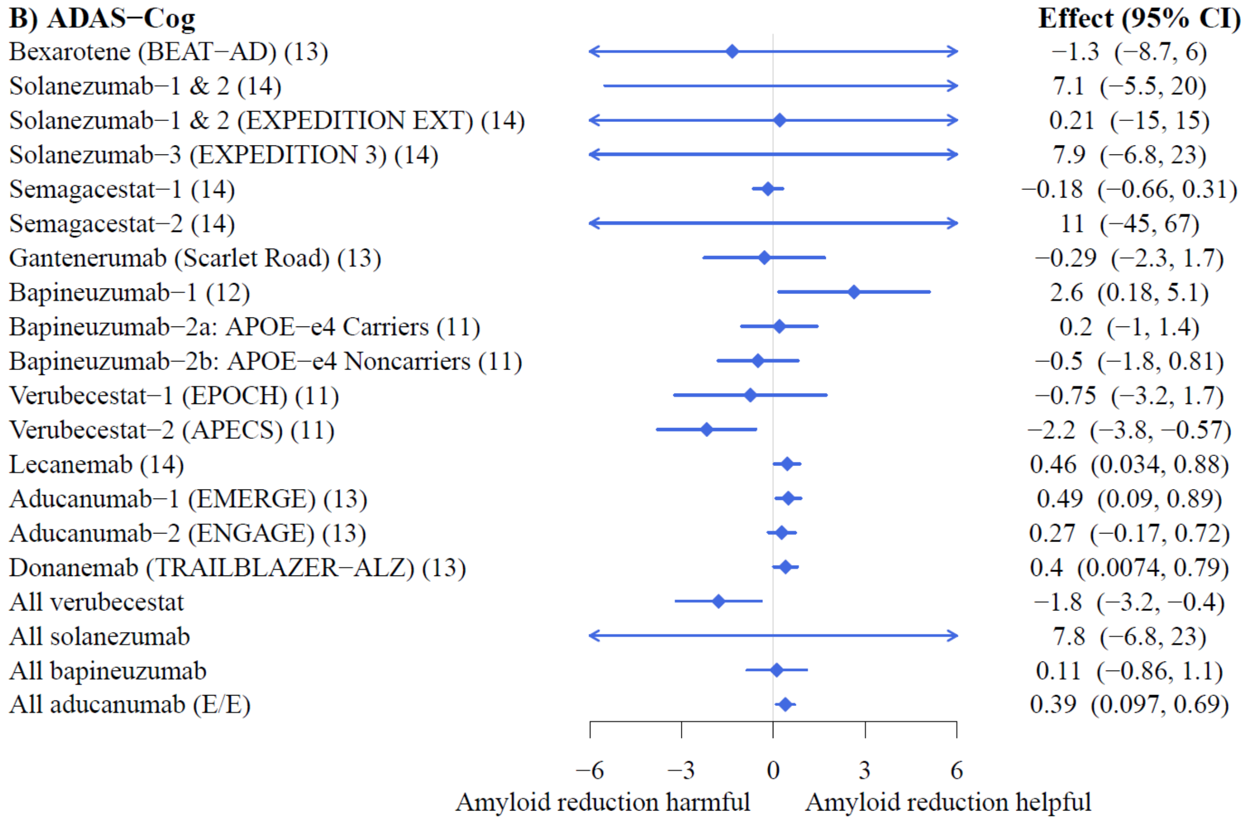

**Figure A1** – C) Trial- and drug-specific estimates representing the effect of each 0.1-unit decrease in brain β-amyloid as measured by PET SUVR on change in MMSE in the clinical trials described alone and for all data from the verubecestat, solanezumab, bapineuzumab, and aducanumab drugs. CI, confidence interval; E/E, EMERGE and ENGAGE; E/E/P, EMERGE, ENGAGE and PRIME; MMSE, Mini-Mental State Examination; PET, positron emission tomography; SUVR, standard uptake value ratio.

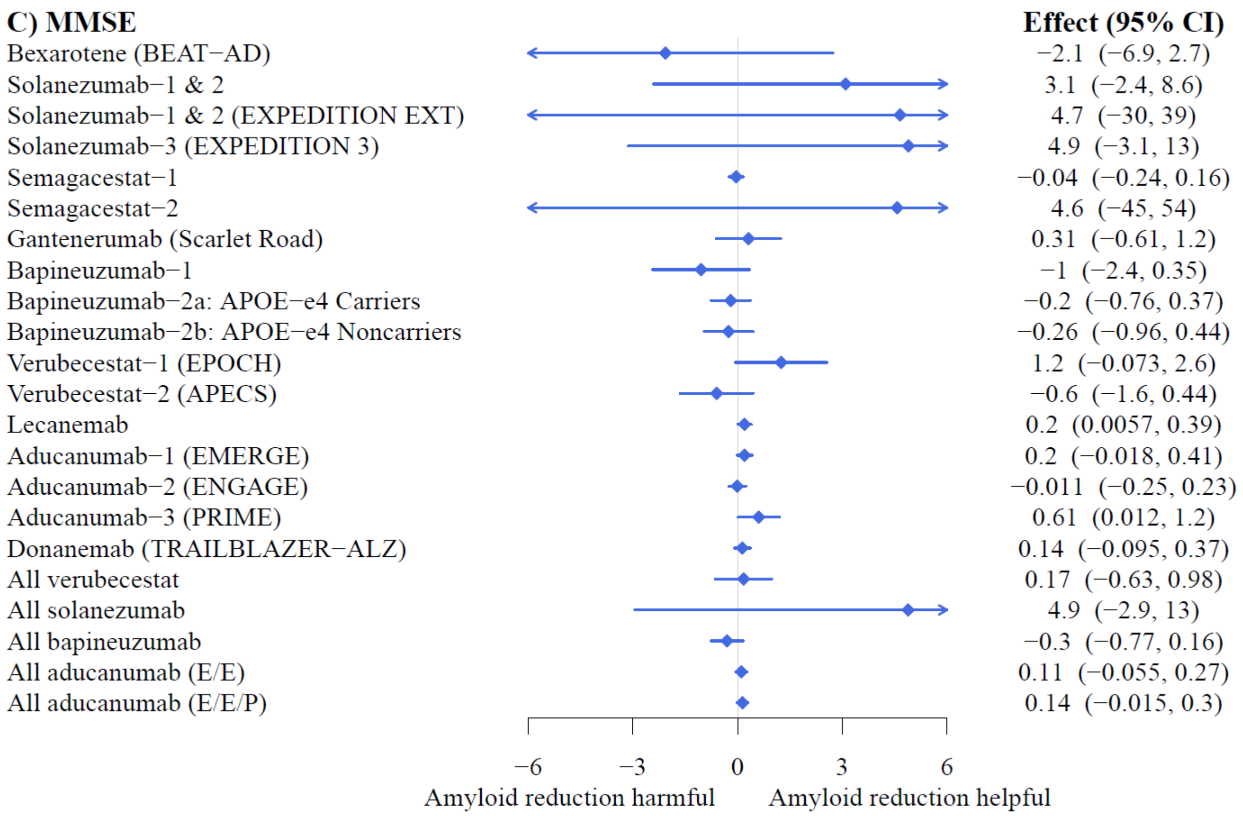

**Figure A2** Pooled estimates from sensitivity analyses representing the effect of each 0.1-unit decrease in brain β-amyloid as measured by PET SUVR on change in A) CDR-SB, B) ADAS-Cog, and C) MMSE. ADAS-Cog, Alzheimer's Disease Assessment Scale–Cognitive Subscale; CDR-SB, Clinical Dementia Rating–Sum of Boxes; CI, confidence interval; MMSE, Mini-Mental State Examination; PET, positron emission tomography; SUVR, standard uptake value ratio.

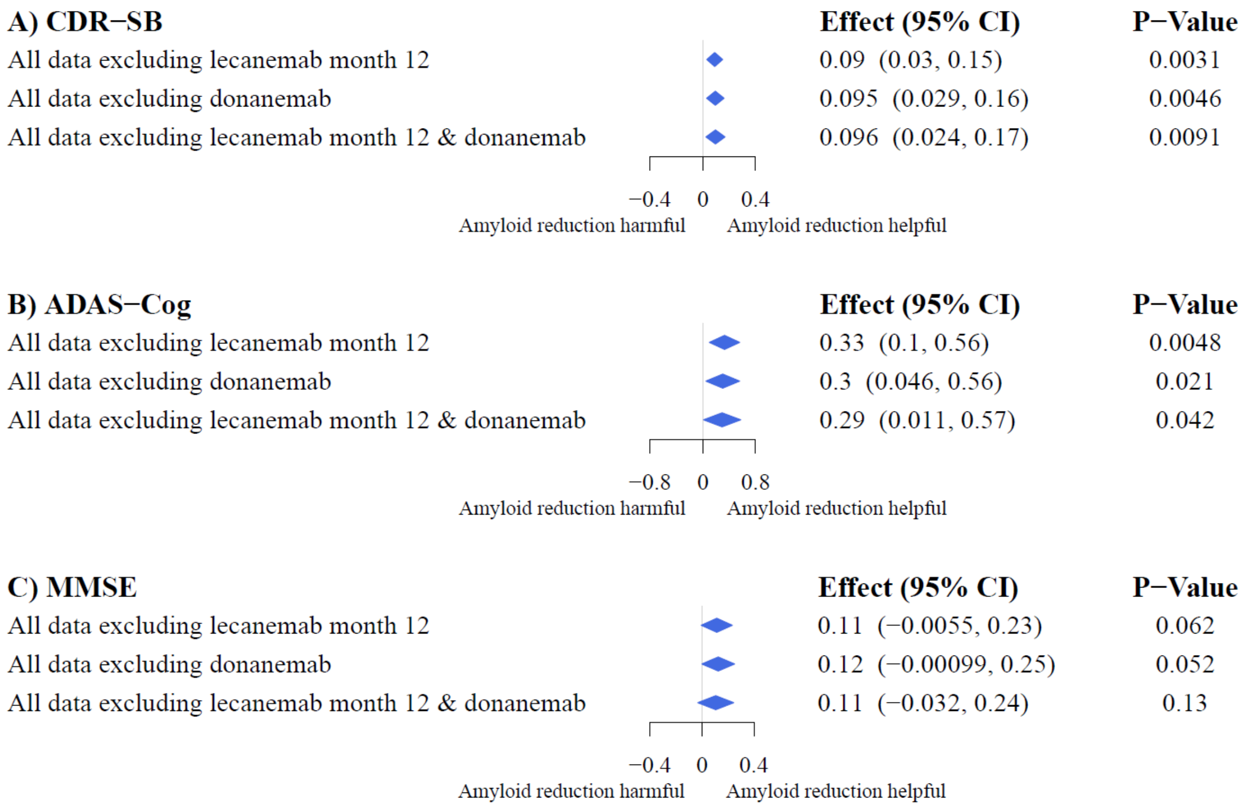

**Appendix 4: Data Source**

| Trial | Trial number | Data source |
|---|---|---|
| Bexarotene (BEAT-AD) | NCT01782742 | Cummings JL, Zhong K, Kinney JW, et al. Double-blind, placebo-controlled, proof-of-concept trial of bexarotene in moderate Alzheimer's disease. *Alzheimers Res Ther*. 2016;8(1):1-9. https://clinicaltrials.gov/ct2/show/results/NCT01782742?term=NCT01782742&draw=2&rank=1 |
| Solanezumab-1&2 (EXPEDITION & EXPEDITION2) | NCT00905372 & NCT00904683 | Siemers ER, Sundell KL, Carlson C, et al. Phase 3 solanezumab trials: secondary outcomes in mild Alzheimer's disease patients. *Alzheimers Dement*. 2016;12(2):110-120. |
| Solanezumab-1&2 Extension (EXPEDITION EXT) | NCT01127633 | https://clinicaltrials.gov/ct2/show/results/NCT01127633?term=NCT01127633&draw=2&rank=1 |
| Solanezumab-3 (EXPEDITION3) | NCT01900665 | https://clinicaltrials.gov/ct2/show/results/NCT01900665?term=Solanezumab&draw=2&rank=2 |
| Semagacestat-1 | NCT00762411 | https://clinicaltrials.gov/ct2/show/results/NCT00762411?term=NCT00762411&draw=2&rank=1 |
| Semagacestat-2 | NCT00594568 | https://clinicaltrials.gov/ct2/show/results/NCT00594568?term=NCT00594568&draw=2&rank=1 |
| Gantenerumab (Scarlet Road) | NCT01224106 | Ostrowitzki S, Lasser RA, Dorflinger E, et al. A phase III randomized trial of gantenerumab in prodromal Alzheimer's disease. *Alzheimers Res Ther*. 2017;9(1):1-5. |
| Bapineuzumab-1 | Registered with EudraCT, number 2004-004120-12; ISRCTN17517446 | Rinne JO, Brooks DJ, Rossor MN, et al. 11C-PiB PET assessment of change in fibrillar amyloid-β load in patients with Alzheimer's disease treated with bapineuzumab: a phase 2, double-blind, placebo-controlled, ascending-dose study. *Lancet Neurol*. 2010;9(4):363-372. |
| Bapineuzumab-2a: APOE-e4 carrier | NCT00575055 | Salloway S, Sperling R, Fox NC, et al. Two phase 3 trials of bapineuzumab in mild-to-moderate Alzheimer's disease. *New Engl J Med*. 2014;370(4):322-333. |
| Bapineuzumab-2b: | NCT00574132 | |

| | | |
|---|---|---|
| APOE-e4 noncarrier | | |
| Verubecestat-1<br>(EPOCH) | NCT01739348 | https://clinicaltrials.gov/ct2/show/results/NCT01739348?term=NCT01739348&draw=2&rank=1 |
| Verubecestat-2<br>(APECS) | NCT01953601 | Egan MF, Kost J, Voss T, et al. Randomized trial of verubecestat for prodromal Alzheimer's disease. *New Engl J Med*. 2019;380(15):1408-1420. |
| Lecanemab* | NCT01767311 | Swanson CJ, Zhang Y, Dhadda S, et al. A randomized, double-blind, phase 2b proof-of-concept clinical trial in early Alzheimer's disease with lecanemab, an anti-Aβ protofibril antibody. *Alzheimers Res Ther*. 2021;13(1):1-4.<br>MMSE data were retrieved from the clinical study report |
| Aducanumab-1<br>(EMERGE) | NCT02484547 | https://clinicaltrials.gov/ct2/show/results/NCT02484547?term=NCT02484547&draw=2&rank=1 |
| Aducanumab-2<br>(ENGAGE) | NCT02477800 | https://clinicaltrials.gov/ct2/show/results/NCT02477800?term=NCT02477800&draw=2&rank=1 |
| Aducanumab-3<br>(PRIME) | NCT01677572 | Data were retrieved from the clinical study report<br><br>Partial data were available in the following publication:<br>Sevigny J, Chiao P, Bussière T, et al. The antibody aducanumab reduces Aβ plaques in Alzheimer's disease. *Nature*. 2016;537(7618):50-56. |
| Donanemab<br>(TRAILBLAZER-ALZ) | NCT03367403 | https://clinicaltrials.gov/ct2/show/results/NCT03367403?term=NCT03367403&draw=2&rank=1<br>Mintun MA, Lo AC, Duggan Evans C, et al. Donanemab in early Alzheimer's disease. *New Engl J Med.* 2021;384(18):1691-1704. |

* Lecanemab trial data were excluded from the "all published data" and "all published antibody data" categories of sensitivity analyses, for all three cognitive endpoints: Alzheimer's Disease Assessment Scale–Cognitive Subscale, Clinical Dementia Rating–Sum of Boxes, and the Mini-Mental State Examination.
